# Development and validation of a cardiovascular diseases risk prediction model for Chinese males (CVDMCM)

**DOI:** 10.1101/2022.02.27.22271588

**Authors:** Ying Shan, Yucong Zhang, Yanping Zhao, Yueqi Lu, Bangwei Chen, Liuqiao Yang, Cong Tan, Yong Bai, Yu Sang, Juehan Liu, Min Jian, Lei Ruan, Cuntai Zhang, Tao Li

## Abstract

**Aims:** Death due to cardiovascular diseases (CVD) increased significantly in China. One way to reduce CVD is to identify people at risk and provide targeted intervention. We aim to develop and validate a CVD risk prediction model for Chinese males (CVDMCM) to help clinicians identify those at risk of CVD and provide targeted intervention.

**Methods and Results:** We conducted a retrospective cohort study of 2331 Chinese males without prior CVD to develop and internally validate the CVDMCM. These participants had a baseline physical examination record (2008-2016) and one revisit record by September 2019. With the full cohort, we used single factor cox regression to examine each candidate predictor adjusted for age. 16 sequential prediction models were built on significant predictors. CVDMCM was selected based on the Akaike information criterion, the area under the ROC curve, and the percentage of variation in outcome values explained by the model (R^2^). This model, the Framingham CVD risk model, and the Wu’s simplified model were all validated by bootstrapping with 1000 repetitions. CVDMCM’s C statistics (0.779, 95% CI: 0.733-0.825), D statistic (4.738, 95% CI: 3.270-6.864), and calibration plot demonstrated that CVDMCM outperformed the other two models.

**Conclusions:** We developed and internally validated CVDMCM, which predicted 4-year CVD risk for Chinese males with a better performance than Framingham CVD model and Wu’s simplified model. In addition, we developed a web calculator for physicians to conveniently generate CVD risk scores and identify those with a higher risk of CVD. We believe CVDMCM had great potential for clinical usage.

## 1 Introduction

According to the World Health Organization, cardiovascular diseases (CVD) are the leading cause of global mortality, accounting for an estimated 17.9 million deaths (31% of all deaths) each year worldwide with an estimated 523 million prevalent CVD cases in 2019 (1). CVD is also the leading cause of death in China, accounting for 45.9% and 43.6% of all deaths in rural and urban China respectively (2). This is more alarming given the proportion of CVD death among all death causes in China used to be 12.8% in 1957 and 35.8% in 1990 (3). China also has the highest CVD burdens internationally, with an estimated 330 million people living with CVD in 2019 (2, 4).

For reducing the CVD burden, one solution is to identify people at higher risk of CVD and offer them appropriate advice for a healthier lifestyle. Thus, numerous prediction models have been developed globally to estimate the risk of CVD, including the Framingham (5-7), SCORE (8), ASSIGN (9), and QRISK models (10, 11). These models were constructed based on data of mainly Caucasian participants rather than Asians, thus, may not be applicable among Chinese. Wu et al. worked out a prediction model based on the USA-PRC Collaborative Study of Cardiovascular and Cardiopulmonary Epidemiology (USA-PRC study) cohort and used the China Multicenter Collaborative Study of Cardiovascular Epidemiology (MUCA) cohort for validation (12). All these models used a 10-year prediction period, which may seem too long for those at risk to take immediate actions. We hypothesized that an estimated risk score in a near future (e.g., within five years) could provide a more powerful warning for the lifestyle change. It was reported that adherence to a healthy lifestyle could lower the CVD burden substantially in the Chinese population (13). According to other research, males were at higher risk of CVD, so we decided to focus on Chinese males first (14). We aimed at developing a CVD risk prediction model for Chinese males (CVDMCM) that could be used by clinicians easily.

We conducted a retrospective cohort study using physical examination data from Tongji Hospital in Wuhan, Hubei Province, China. Participants were Chinese males who underwent annual physical examinations from 2008 to 2016 and had at least one revisit record by September 2019. We collected data of 2331 30 to 74-year-old Chinese males for the analysis. The main CVD outcomes included coronary heart disease (CHD) and ischemic stroke. We developed and internally validated a 4-year CVD risk prediction model among Chinese males. The full cohort was used as the derivation cohort and internal model validation was carried out by bootstrapping with 1000 repetitions. CVDMCM was internally validated by bootstrap resampling and compared with established models, namely the Framingham CVD risk model and Wu’s simplified prediction model (12, 15).

## 2 Methods

### 2.1 Study design

We conducted a retrospective cohort study to develop CVDMCM. The participants were Chinese males from the general population going through annual physical examination at Tongji Hospital in Wuhan, Hubei Province in central China. The physical examination department of Tongji Hospital is the biggest physical examination center in Wuhan.

Inclusion and exclusion criteria: we included Chinese males aged 30-74 years old who took physical examinations at Tongji Hospital between 2008 and 2016 and had at least one revisit record by September 2019. The baseline record was defined as the first time a participant took the examination between 2008 and 2016. At baseline, participants with the following conditions were excluded: those with a CHD and ischemic stroke, other heart diseases (i.e., rheumatic heart disease), a malignant tumor, or a history of liver or kidney failure. If the participants had more than one revisit record and no outcome event occurred by September 2019, we used their last revisit date and their free-of-CVD status. If there was an event by September 2019, we used their CVD event reporting date and CVD event status.

This study followed the Transparent Reporting of a multivariable prediction model for Individual Prognosis or Diagnosis (TRIPOD) statement (16). The study was approved by the Medical Ethics Committee at Tongji Medical College, Huazhong University of Science and Technology who waived the written informed consents (TJ-IRB20191215). Participants identified with CVD would be recommended to go to see the doctor. No intervention was provided in the physical examination center.

### 2.2 Potential predictors

Based on prior knowledge, we included 17 potential predictors, including lifestyle characteristics, clinical measurements, and medical history indicators collected by trained doctors at the physical examination center of Tongji Hospital through standardized in-person interviews (17). The potential predictors included both classic ones, such as those in the Framingham CVD model, and more modern ones, such as ankle brachial index (ABI) and brachial-ankle pulse wave velocity (baPWV). Specific measuring protocols are listed in the Supplementary Methods.

### 2.3 Outcomes

The CVD outcomes included CHD and ischemic stroke. CHD was diagnosed according to symptoms (mainly angina) and electrocardiography or coronary angiography. Patients who self-reported having coronary heart disease, coronary artery bypass grafting, coronary stent implantation, percutaneous coronary intervention, or percutaneous transluminal coronary angioplasty in the follow-up visits were also considered to have CHD. Ischemic stroke was diagnosed according to symptoms and cerebral infarction and confirmed by computed tomography or magnetic resonance imaging.

### 2.4 Sample size

We calculated the minimum sample size required for the prediction model according to Riley’s guidance (18). By treating CVD occurrences as time-to-event outcomes, sample size calculations are provided to (S1) estimate the overall outcome proportion with precision in follow-up, (S2) target a shrinkage factor of 0.9, and (S3) target small optimism of 0.05 in the apparent R^2^. Based on the 3 criteria, the sample size in the cohort is expected to be sufficient. The details are shown below:

We used R 4.0.3 package ‘pmsampsize’ for criteria S1, S2, and S3 where the anticipated R^2^ value was assumed to be 0.25, according to existing CVD risk prediction models shown in Siontis et al., with up to 20 parameters (19). Siontis et al. illustrated R^2^ statistics of 12 existing CVD risk prediction models, and all the R^2^ statistics are higher than 0.25 (19). Therefore, we made a conservative choice of R^2^ of 0.20. For the convenience of clinical application, the number of parameters in the final model tends to be no more than 10. Again, to be conservative, we set the model with up to 20 parameters. The mean follow-up and the overall event rate were calculated in our study cohort. The timepoint of interest for prediction using the newly developed CVDMCM was 4 years.

### 2.5 Model development

First, physical examination records which met our inclusion criteria were exported from the electronic medical record system of Tongji Hospital. We then excluded the participants who met the exclusion criteria. A retrospective cohort was established afterwards. To deal with the missing data, we applied multiple imputations to the raw data (20). The imputations were implemented with Multivariate Imputation by Chained Equations (MICE) package in R (21). Then, we examined the baseline characteristics for the cohorts, summarized categorical data with frequencies and percentages, as well as summarized continuous data with means and standard deviations (if normally distributed) or medians and interquartile ranges (if not normally distributed).

We constructed CVDMCM with the cohort using Cox proportional hazards regression. The quadratic terms of predictors, such as systolic blood pressure (SBP), diastolic blood pressure (DBP), body mass index (BMI), and glycated hemoglobin A1c (HbA1c) were considered. The interaction effects between age and the predictors were also assessed because the effects of some factors may change with age. We first fitted cox regression models of each predictor along with age and collected the statistically significant predictors (*P* < 0.05). Then, we constructed sequential models with those significant predictors along with age. We compared those sequential models with the Akaike information criterion (AIC), area under the ROC curve (AUC), and R^2^, where AUC was calculated under the bootstrap strategy of 1000 replicates. We selected the most fitted model based on AIC, as well as considered the other indicators. We assessed the proportional hazards assumption of each predictor by examining the plots of the scaled Schoenfeld residuals against time. Any non-random pattern in the plots suggested a violation of the proportional hazard’s assumptions, in which we attempted the log transformation of the covariates.

We then defined the high-risk individuals as having a risk higher than the age-standardized CVD prevalence rate of Chinese males in literature (22, 23). This would provide physicians with a cutoff to inform those individuals with high CVD risk score to change their lifestyle, take more physical activities, and pay close attention to their cardiovascular health.

### 2.6 Validation and model performance

We applied bootstrap resampling with 1000 replicates to evaluate CVDMCM. The performance of CVDMCM was assessed via calibration and discrimination and compared with the Framingham CVD risk model and the simplified model in Wu et al. (12, 15). Calibration demonstrated model performance concerning agreement between predicted risk and observed risk and was reported using a calibration plot (24). We generated calibration plots based on the predicted and 4-year observed CVD risks. Discrimination, which is defined as the model’s ability to distinguish CVD and non-CVD, will be measured via calculation of Harrell’s C statistic and D statistic. Harrell’s C statistic is known as the concordance index. Though it works like AUC, Harrell’s C statistic is better than AUC as it takes into account the censoring pattern of the participants (25). For the model to be acceptable, its Harrell’s C statistic should be at least 0.7. If its Harrell’s C statistic is over 0.9, it shows that the model has excellent predictive power (26). The D statistic is also a measure of discrimination. A higher value of D statistic indicates better discrimination (27).

## 3 Results

We initially included 2470 participants who met the inclusion criteria. From those 2470 participants, we excluded 139 participants who met the exclusion criteria. As the result, 2331 individuals remained in this study. The study flow diagram could be found in Figure 1. The participants, both with and without CVD events, were followed for a median of 4.0 years (Table 1). The mean baseline age of the participants was 52.3 years old with a standard deviation (SD) of 9.2. The baseline age of the participants who had CVD events during the follow-up was 60.8 (SD: 7.2) years old, while the baseline age of those without CVD events was 51.8 (SD: 9.1) years old. The incidence rate of CVD per 1000 person-years was 13.16 (95% CI: 11.00-15.63).

**Figure 1.**
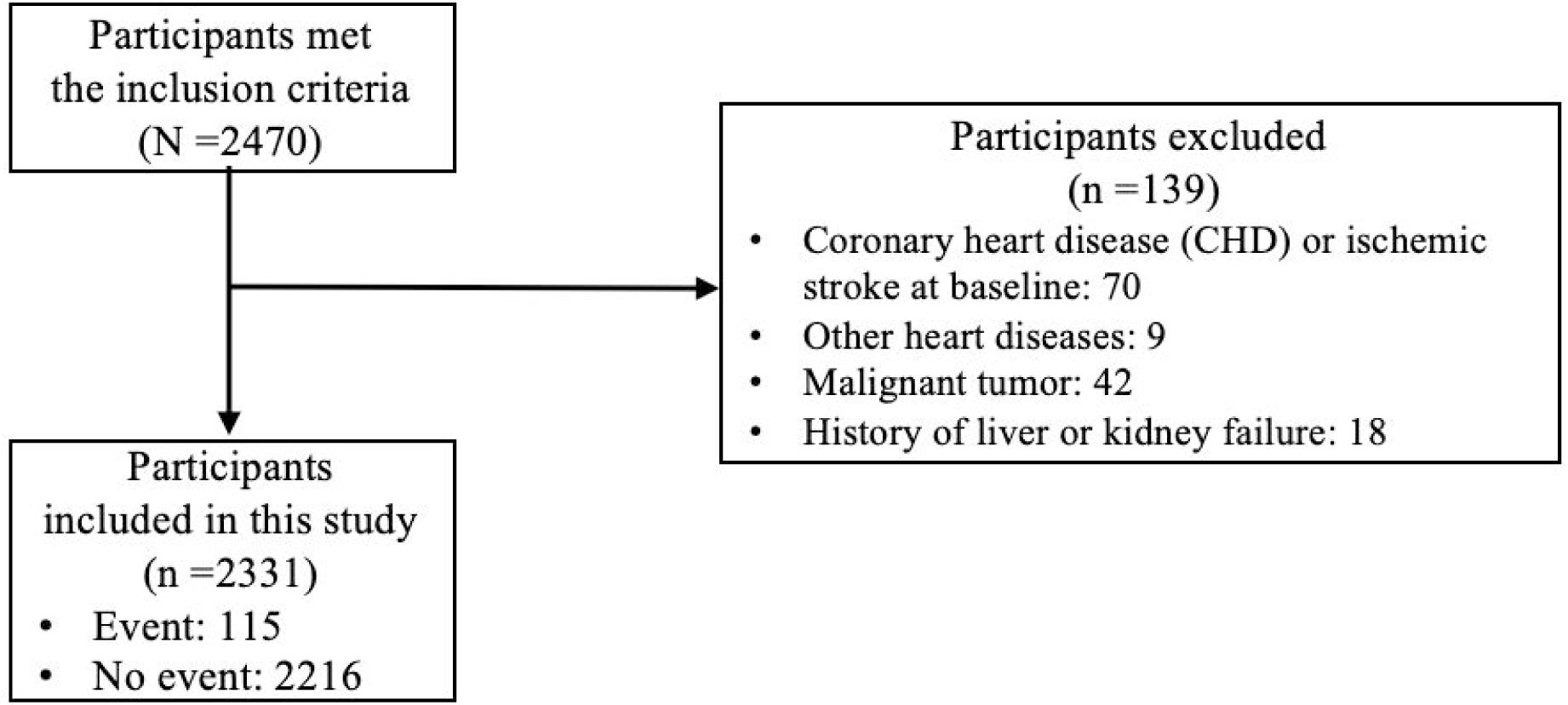
Study flow diagram.

**Table 1.**
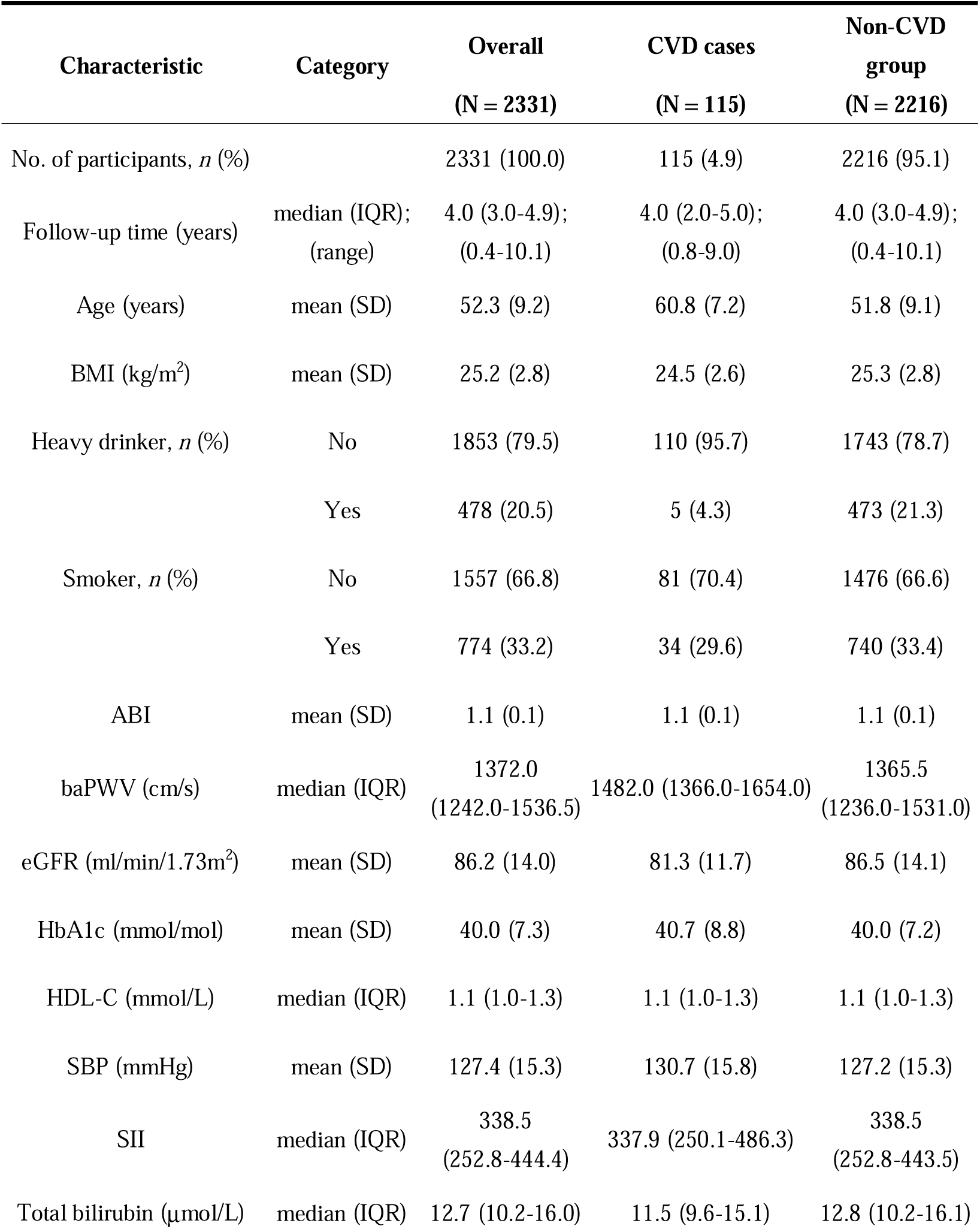
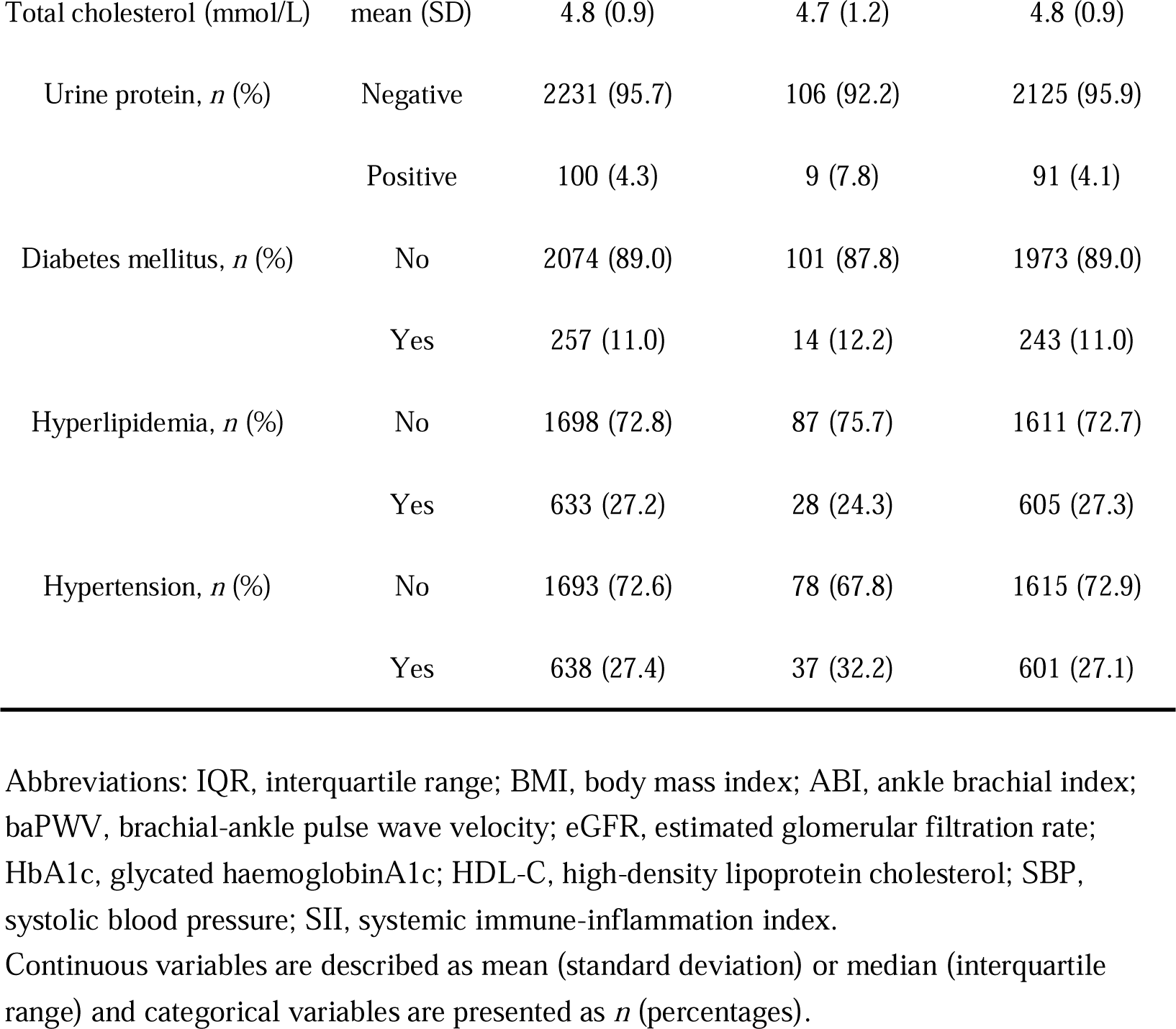
Baseline characteristics.

Then, we calculated the minimum sample size required by using ‘pmsampsize’ R package with the parameters of R^2^ value assumed to be 0.20 with up to 20 parameters. The mean follow-up of 4.23 years and the overall event rate of 0.049 were obtained in our study cohort. The timepoint of interest was 4 years. The calculated minimum sample size required was 796, which indicated that the sample size in the cohort of 2331 was sufficient.

CVDMCM was derived using the potential predictors, along with the interaction terms and the quadratic terms by single factor Cox regression adjusting for age. Five predictors were significant. Along with age, the significant predictors were used to construct 16 sequential prediction models. The plots of the scaled Schoenfeld residuals against time suggested that all the 16 models fulfilled the proportional hazards assumption. Table 2 compared the 16 sequential models with AIC, AUC, and R^2^. Among the sequential models, model 15 had the smallest AIC value, a relatively big AUC, and the biggest R^2^ values. Thus, we chose model 15 as our final CVDMCM and compared it with the Framingham CVD risk model and Wu’s simplified model. Figure S1 showed the Schoenfeld individual test of each predictor in model 15 (figures not shown for the other models). In model 15, predictors were age, smoking status, log-transformed total bilirubin, ABI, SBP, and the interaction of age and SBP. The model is shown below:

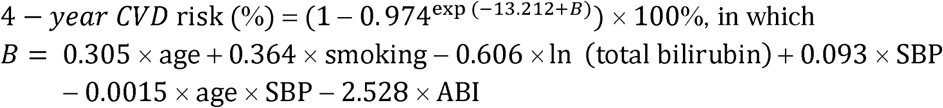

We calculated the age-standardized CVD prevalence rate of Chinese males according to Liu et al., which was 3.14% (23). An individual with an estimated risk higher than 3.14% is defined as a high-risk individual. In the cohort, 1053 out of 2331 individuals would be classified in the high-risk group.

**Table 2.** The hazard ratio of the predictors in the 16 sequential prediction models. The AIC, AUC and R^2^ of the 16 sequential models.

| Predictors | Model 1 | Model 2 | Model 3 | Model 4 | Model 5 | Model 6 | Model 7 | Model 8 | Model 9 | Model 10 | Model 11 | Model 12 | Model 13 | Model 14 | Model 15 | Model 16 |
| --- | --- | --- | --- | --- | --- | --- | --- | --- | --- | --- | --- | --- | --- | --- | --- | --- |
| Age | 1.10 | 1.10 | 1.10 | 1.10 | 1.34 | 1.10 | 1.11 | 1.33 | 1.10 | 1.33 | 1.36 | 1.11 | 1.33 | 1.36 | 1.36 |  |
| Smoking status |  | 1.47 |  |  |  | 1.44 | 1.52 | 1.44 |  |  |  | 1.49 | 1.40 |  | 1.44 | 1.47 |
| Log total bilirubin |  |  | 0.51 |  |  | 0.53 |  |  | 0.52 | 0.52 |  | 0.54 | 0.53 | 0.53 | 0.55 | 0.53 |
| ABI |  |  |  | 0.01 |  |  | 0.08 |  | 0.11 |  | 0.08 | 0.09 |  | 0.09 | 0.08 | 0.11 |
| SBP |  |  |  |  | 1.09 |  |  | 1.09 |  | 1.09 | 1.10 |  | 1.09 | 1.10 | 1.10 | 0.95 |
| Age x SBP |  |  |  |  | 0.99 |  |  | 0.99 |  | 0.99 | 0.99 |  | 0.99 | 0.99 | 0.99 | 1.00 |
| AIC | 1715.69 | 1714.31 | 1710.74 | 1713.18 | 1714.71 | 1709.76 | 1711.24 | 1713.79 | 1708.68 | 1710.02 | 1711.46 | 1707.21 | 1709.49 | 1707.12 | 1706.22 | 1715.48 |
| AUC | 0.78 | 0.79 | 0.78 | 0.79 | 0.78 | 0.78 | 0.79 | 0.79 | 0.79 | 0.78 | 0.79 | 0.79 | 0.78 | 0.79 | 0.79 | 0.78 |
| R<sup>2</sup> | 0.36 | 0.38 | 0.39 | 0.38 | 0.38 | 0.4 | 0.39 | 0.39 | 0.40 | 0.41 | 0.40 | 0.42 | 0.42 | 0.42 | 0.43 | 0.39 |

We then internally validated the CVDMCM and compared its performance with Framingham and Wu’s model by applying a bootstrap resampling approach with 1000 replicates. We first compared the predictors of CVDMCM, the Framingham CVD risk model, and Wu’s model. As shown in Table 3, the predictors used in all three models included age, smoking status, and SBP. Compared with Framingham and Wu’s models, CVDMCM included more modern predictors, such as ABI and Total bilirubin. We then compared the calibration of the 3 models. As shown in Figure 2, CVDMCM demonstrated higher agreement between 4-year predicted risk and observed risk, which indicated better calibration than the Framingham and Wu’s simplified model. Finally, discrimination was assessed via Harrell’s C statistic and D statistic. As shown in Table 4, CVDMCM performed better than the Framingham CVD risk model and Wu’s simplified model with higher Harrell’s C statistic of 0.779 (CI: 0.733-0.825), and D statistic of 4.738 (CI: 3.270-6.864). The Harrell’s C statistic of CVDMCM was significantly higher than that of Framingham CVD risk model and that of Wu’s simplified model with both of the one-sided *P*-values smaller than 0.001.

**Table 3.**
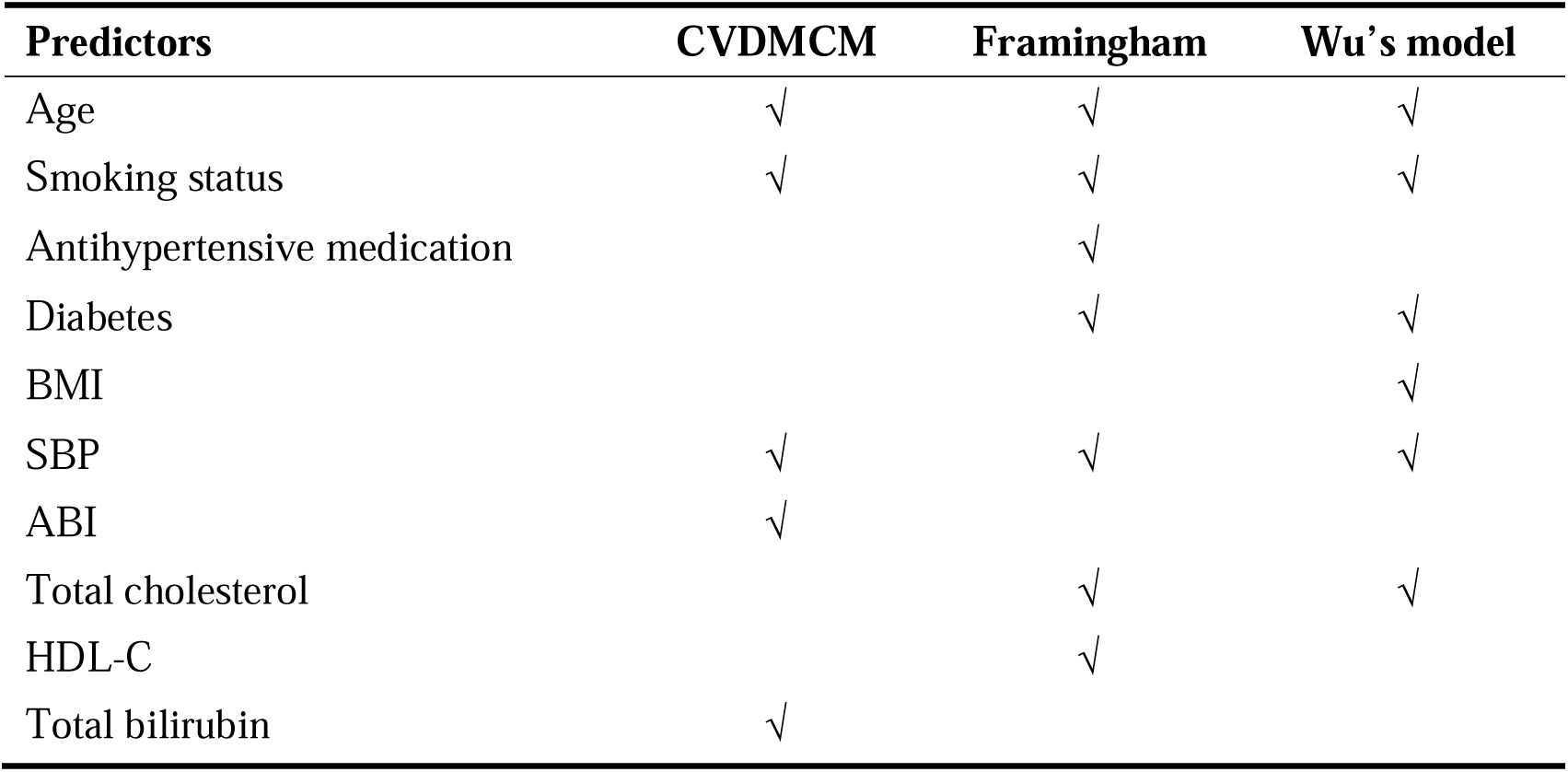
Predictors used in CVDMCM, the Framingham and Wu’s model for males.

**Figure 2.**
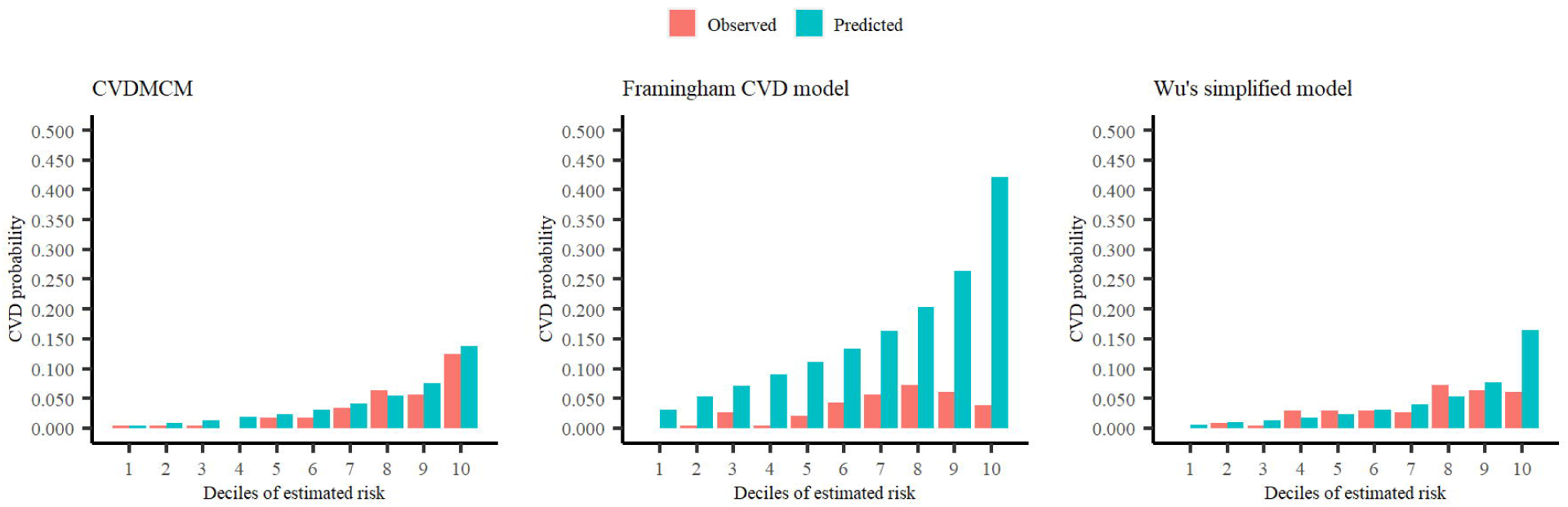
Calibration plots of CVDMCM, the Framingham CVD risk model, and Wu’s simplified model for observed and predicted 4-year risks of CVD using a validation dataset.

**Table 4.**
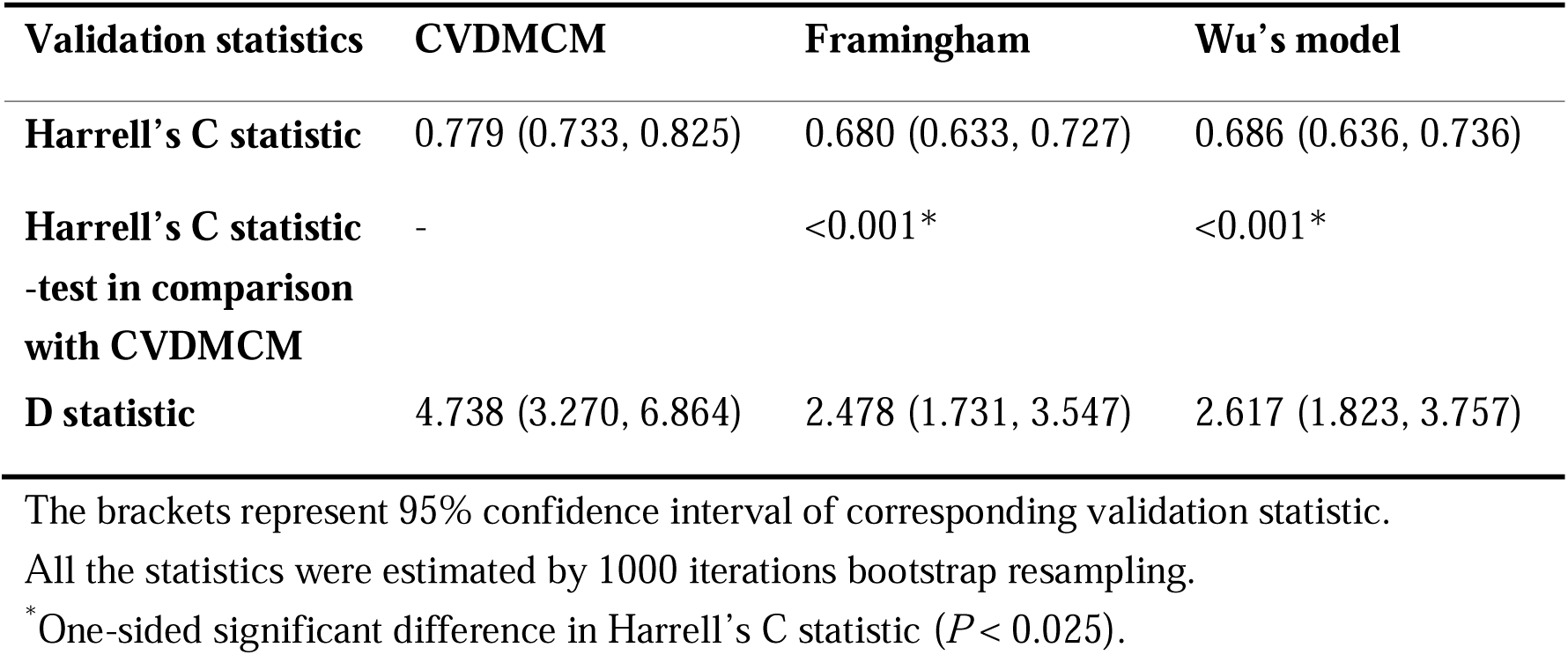
Performance of CVDMCM, the Framingham and Wu’s model in the validation cohort for predicting 4-year risk of CVD.

For an easier application of CVDMCM among the clinicians, we provided one example of an individual Mr. X’s 4-year CVD risk. A 54-year-old Chinese man, Mr. X, who is a non-smoker with a total bilirubin of 12.30 μmol/L, ABI of 3.39, and SBP of 130mmHg, has a 4-year CVD risk of 2.39%. The calculation is shown below:

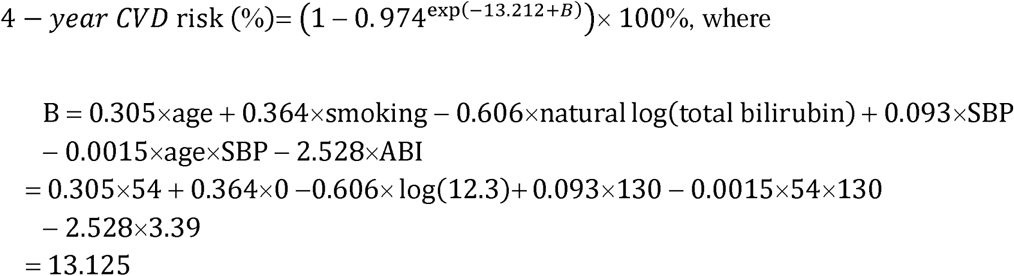

So, the 4-year CVD risk is

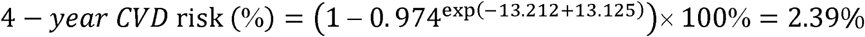

Since his CVD risk is lower than 3.14%, he would be classified in the low-risk group. If someone has a CVD risk calculated higher than 3.14%, he would be classified in the high-risk group, and the physicians taking the physical examination could advise him for a healthier lifestyle. To make the CVDMCM more convenient for the physicians, we have developed an online calculator calCVDrisk (https://ctan2020.github.io/-calCVDrisk-/) for the physicians to simply enter the parameters obtained from the physical examinations, and the calculator would report the score for the physicians to assess the CVD risks.

## 4 Discussion

Through this retrospective cohort study, we developed and validated a CVD risk prediction model CVDMCM. Unlike models with tens of predictors, CVDMCM had only five predictors (age, smoking status, total bilirubin, ABI, and SBP) and demonstrated a better performance than that of the Framingham CVD risk model and Wu’s simplified model in the internal validation by bootstrap resampling as found in Harrell’s C statistic, D statistic, and calibration plot (12, 15). We believe that CVDMCM had great potential for clinical usage.

There were several strengths of this study. First, we targeted a high-risk population, namely Chinese males. Given that CVD is the leading cause of death in China accounting for over 43.6% of all deaths in China, which was an increase from 12.8% in 1957 and 35.8% in 1990, immediate action is needed to change the increasing trend (3). Among the Chinese population, Chinese males were at higher risk of CVD. Thus, we decided to work on a model for the Chinese males in this study (14). Secondly, we used a 4-year prediction model. Previously, CVD prediction models were often 10-year risks, which may seem too long for those at risk to take immediate actions (8, 12, 15, 28). We hypothesized that an estimated risk score in four years could provide a more powerful early warning for the lifestyle change. Thirdly, for an easier application of our finding, we have developed an online calculator calCVDrisk for physicians to assess the CVD risk scores among those taking physical examinations.

We admit that there are some limitations to this study. First, there is inevitably selection bias by choosing the participants taking physical examinations. They might take better care of their health than those who do not take regular physical examinations. A model to identify those at higher risk of CVD with appropriate lifestyle change advice might work with this population, as they pay more attention to their health and are more likely to follow the doctor’s advice and modify their lifestyles than those who do not regularly take physical examinations. If the model works for this population, we could test it on other populations later. Secondly, the data obtained from electronic medical records from only one hospital may induce selection bias because it may not represent the general population of China. However, as Tongji Hospital locates in the city of Wuhan, which is considered as the geographic midpoint of China, the location might well represent the majority of Chinese cities with patients coming from all over China. The electronic medical records are from the physical examination department of Tongji Hospital, which is the biggest physical examination center in Wuhan where various groups of people choose to go to take the medical examination. Thus, the data may well represent Chinese males. Thirdly, the study has a bias of a loss to follow-up, which should be improved in future studies.

Compared with the classic CVD models, such as the Framingham (5-7), SCORE (8), ASSIGN (9), and QRISK models (10, 11), CVDMCM was developed using Chinese males’ data, which is more applicable in China. A previous study reported that CVD models should better be developed among specific populations (29). Although Wu’s model is a very well designed one among the Chinese, it was published 15 years ago, thus, did not include more modern predictors such as ABI and baPWV. ABI was found as one of the 5 predictors in CVDMCM. ABI has become widely used in modern clinical practice and has been reported to be a sensitive measurement of CVD risk. L. Alves-Cabratosa et al. reported that low ABI would increase the risk of acute myocardial infarction and ischemic stroke among asymptomatic people, as well as people with diabetes and previously-diagnosed CVD (30). A.Velescu et al. showed that adding ABI to the Framingham CVD risk model would improve the capacity of predicting CVD events in northeastern Spain population (31).

To better utilize CVDMCM, we developed an online calculator calCVDrisk for physicians to calculate the CVD risk of a Chinese male when taking the physical examination. We expect that CVDMCM will be promising in the application as it is a simple model with only 5 predictors. A previous study has shown that physicians were more willing to use simpler models in clinical practice than more complex ones when they had comparable performance (32). The estimated 4-year CVD risk will let people be alert to their cardiovascular health. A prospective cohort is under plan to use CVDMCM to detect those at higher risk of CVD to further validate the model at the physical examination department of Tongji Hospital. If the results are promising, we would advocate this application in major physical examination departments in China.

We believe that the CVDMCM could be helpful for medical doctors in the physical examination departments in China to select those at higher risks of CVD in 4 years. For health care providers, the integration of a better CVD risk prediction model in their workflow would enable them to track the health statuses of those with high-risk scores so that doctors could provide advice for lifestyle changes and potentially help reduce CVD risks in those patients.

## 5 Conclusion

We conducted a retrospective cohort study for developing and validating our new model, CVDMCM. CVDMCM predicted 4-year CVD risk with a better performance than that of the Framingham CVD model and with fewer predictors than those involved in the Wu’s simplified model. Thus, this model could be helpful in clinical practices to detect patients at higher risk of CVD, providing an appropriate feedback for patients’ health. We believe that CVDMCM had great potential for clinical usage.

## Data Availability

All data produced in the present study are available upon reasonable request to the authors.

## 6 Supplementary materials

Supplementary materials are available at Frontiers in Cardiovascular Medicine online.

## 8 Author Contribution Statement

Y.Shan, Y.Zhao, and T.Li conceived the project, designed the research study. L.Ruan, C.Zhang and Y.Sang collected the data of the cohort. Y.Shan, Y.Lu, B.Chen, L. Yang analyzed data. Y.Shan, Y.Zhao, Y.Lu, B.Chen, L.Yang, Y.Bai, J.Liu and M.Jian wrote the manuscript. The co-first authors each made important contributions to different aspects of the project, including designing the project, collecting the data and analyzing the data. The final order was discussed and agreed upon by placing authors who contributed to project design and data analysis higher on the list.

## 9 Acknowledgements

We would like to thank the general practitioners, especially those of the Department of Geriatrics of Tongji Hospital, Tongji Medical College, Huazhong University of Science and Technology for their dedication, commitment, and contribution.

## 10 Data Availability statement

The data underlying this article will be shared at reasonable request to the corresponding author.

## References

1. World Health Organization. Cardiovascular diseases (CVDs). Available from: https://www.who.int/news-room/fact-sheets/detail/cardiovascular-diseases-(cvds)

2. The Writing Committee of the Report on Cardiovascular Health and Diseases in China. Report on Cardiovascular Health and Diseases in China 2019: an Updated Summary. Chinese Circulation Journal. 2020;35(09)

3. Khor GL. Cardiovascular epidemiology in the Asia-Pacific region. Asia Pac J Clin Nutr. 2001;10(2):76–80.doi: 10.1111/j.1440-6047.2001.00230.x

4. Zhao D, Liu J, Wang M, Zhang X, Zhou M. Epidemiology of cardiovascular disease in China: current features and implications. Nat Rev Cardiol. 2019;16(4):203–12.doi: 10.1038/s41569-018-0119-4

5. Anderson KM, Odell PM, Wilson PW, Kannel WB. Cardiovascular disease risk profiles. Am Heart J. 1991;121(1 Pt 2):293–8.doi: 10.1016/0002-8703(91)90861-b

6. Anderson KM, Wilson PW, Odell PM, Kannel WB. An updated coronary risk profile. A statement for health professionals. Circulation. 1991;83(1):356–62.doi: 10.1161/01.cir.83.1.356

7. Wilson PW, D’Agostino RB, Levy D, Belanger AM, Silbershatz H, Kannel WB. Prediction of coronary heart disease using risk factor categories. Circulation. 1998;97(18):1837–47.doi: 10.1161/01.cir.97.18.1837

8. Conroy RM, Pyorala K, Fitzgerald AP, Sans S, Menotti A, De Backer G, et al. Estimation of ten-year risk of fatal cardiovascular disease in Europe: the SCORE project. Eur Heart J. 2003;24(11):987–1003.doi: 10.1016/s0195-668x(03)00114-3

9. Woodward M, Brindle P, Tunstall-Pedoe H, estimation Sgor. Adding social deprivation and family history to cardiovascular risk assessment: the ASSIGN score from the Scottish Heart Health Extended Cohort (SHHEC). Heart. 2007;93(2):172–6.doi: 10.1136/hrt.2006.108167

10. Hippisley-Cox J, Coupland C, Robson J, Brindle P. Derivation, validation, and evaluation of a new QRISK model to estimate lifetime risk of cardiovascular disease: cohort study using QResearch database. BMJ. 2010;341:c6624.doi: 10.1136/bmj.c6624

11. Hippisley-Cox J, Coupland C, Vinogradova Y, Robson J, May M, Brindle P. Derivation and validation of QRISK, a new cardiovascular disease risk score for the United Kingdom: prospective open cohort study. BMJ. 2007;335(7611):136.doi: 10.1136/bmj.39261.471806.55

12. Wu Y, Liu X, Li X, Li Y, Zhao L, Chen Z, et al. Estimation of 10-year risk of fatal and nonfatal ischemic cardiovascular diseases in Chinese adults. Circulation. 2006;114(21):2217–25.doi: 10.1161/CIRCULATIONAHA.105.607499

13. Lv J, Yu C, Guo Y, Bian Z, Yang L, Chen Y, et al. Adherence to Healthy Lifestyle and Cardiovascular Diseases in the Chinese Population. J Am Coll Cardiol. 2017;69(9):1116–25.doi: 10.1016/j.jacc.2016.11.076

14. Yang ZJ, Liu J, Ge JP, Chen L, Zhao ZG, Yang WY, et al. Prevalence of cardiovascular disease risk factor in the Chinese population: the 2007-2008 China National Diabetes and Metabolic Disorders Study. Eur Heart J. 2012;33(2):213–20.doi: 10.1093/eurheartj/ehr205

15. D’Agostino RB, Sr., Vasan RS, Pencina MJ, Wolf PA, Cobain M, Massaro JM, et al. General cardiovascular risk profile for use in primary care: the Framingham Heart Study. Circulation. 2008;117(6):743–53.doi: 10.1161/CIRCULATIONAHA.107.699579

16. Collins GS, Reitsma JB, Altman DG, Moons KG. Transparent reporting of a multivariable prediction model for individual prognosis or diagnosis (TRIPOD): the TRIPOD statement. BMJ. 2015;350:g7594.doi: 10.1136/bmj.g7594

17. Joseph Loscalzo M-H. Harrison’s Cardiovascular Medicine, 3/E2016.

18. Riley RD, Ensor J, Snell KIE, Harrell FE Jr.,, Martin GP, Reitsma JB, et al. Calculating the sample size required for developing a clinical prediction model. BMJ. 2020;368:m441.doi: 10.1136/bmj.m441

19. Siontis GC, Tzoulaki I, Siontis KC, Ioannidis JP. Comparisons of established risk prediction models for cardiovascular disease: systematic review. BMJ. 2012;344:e3318.doi: 10.1136/bmj.e3318

20. Yucel RM. Multiple imputation inference for multivariate multilevel continuous data with ignorable non-response. Philos Trans A Math Phys Eng Sci. 2008;366(1874):2389–403.doi: 10.1098/rsta.2008.0038

21. Buuren S, Groothuis-Oudshoorn C. MICE: Multivariate Imputation by Chained Equations in R. Journal of Statistical Software. 2011;45.doi: 10.18637/jss.v045.i03

22. Shan Y, Tromp G, Kuivaniemi H, Smelser DT, Verma SS, Ritchie MD, et al. Genetic risk models: Influence of model size on risk estimates and precision. Genet Epidemiol. 2017;41(4):282–96.doi: 10.1002/gepi.22035

23. Liu S, Li Y, Zeng X, Wang H, Yin P, Wang L, et al. Burden of Cardiovascular Diseases in China, 1990-2016: Findings From the 2016 Global Burden of Disease Study. JAMA Cardiol. 2019;4(4):342–52.doi: 10.1001/jamacardio.2019.0295

24. Steyerberg EW, Vickers AJ, Cook NR, Gerds T, Gonen M, Obuchowski N, et al. Assessing the performance of prediction models: a framework for traditional and novel measures. Epidemiology. 2010;21(1):128–38.doi: 10.1097/EDE.0b013e3181c30fb2

25. Wan EYF, Fong DYT, Fung CSC, Yu EYT, Chin WY, Chan AKC, et al. Development of a cardiovascular diseases risk prediction model and tools for Chinese patients with type 2 diabetes mellitus: A population-based retrospective cohort study. Diabetes Obes Metab. 2018;20(2):309–18.doi: 10.1111/dom.13066

26. Swets JA. Measuring the accuracy of diagnostic systems. Science. 1988;240(4857):1285–93.doi: 10.1126/science.3287615

27. Hippisley-Cox J, Coupland C, Brindle P. Development and validation of QRISK3 risk prediction algorithms to estimate future risk of cardiovascular disease: prospective cohort study. BMJ. 2017;357:j2099.doi: 10.1136/bmj.j2099

28. Yang X, Li J, Hu D, Chen J, Li Y, Huang J, et al. Predicting the 10-Year Risks of Atherosclerotic Cardiovascular Disease in Chinese Population: The China-PAR Project (Prediction for ASCVD Risk in China). Circulation. 2016;134(19):1430–40.doi: 10.1161/CIRCULATIONAHA.116.022367

29. Damen JA, Pajouheshnia R, Heus P, Moons KGM, Reitsma JB, Scholten R, et al. Performance of the Framingham risk models and pooled cohort equations for predicting 10-year risk of cardiovascular disease: a systematic review and meta-analysis. BMC Med. 2019;17(1):109.doi: 10.1186/s12916-019-1340-7

30. Alves-Cabratosa L, Garcia-Gil M, Comas-Cufi M, Blanch J, Ponjoan A, Marti-Lluch R, et al. Role of Low Ankle-Brachial Index in Cardiovascular and Mortality Risk Compared with Major Risk Conditions. J Clin Med. 2019;8(6).doi: 10.3390/jcm8060870

31. Velescu A, Clara A, Penafiel J, Ramos R, Marti R, Grau M, et al. Adding low ankle brachial index to classical risk factors improves the prediction of major cardiovascular events. The REGICOR study. Atherosclerosis. 2015;241(2):357–63.doi: 10.1016/j.atherosclerosis.2015.05.017

32. Brand EC, Elias SG, Minderhoud IM, van der Veen JJ, Baert FJ, Laharie D, et al. Systematic Review and External Validation of Prediction Models Based on Symptoms and Biomarkers for Identifying Endoscopic Activity in Crohn’s Disease. Clin Gastroenterol Hepatol. 2020;18(8):1704–18.doi: 10.1016/j.cgh.2019.12.014

