## Supplementary material for "Development and validation of a cardiovascular diseases risk prediction model for Chinese males (CVDMCM)": Figure S1

**Supplementary Figures**

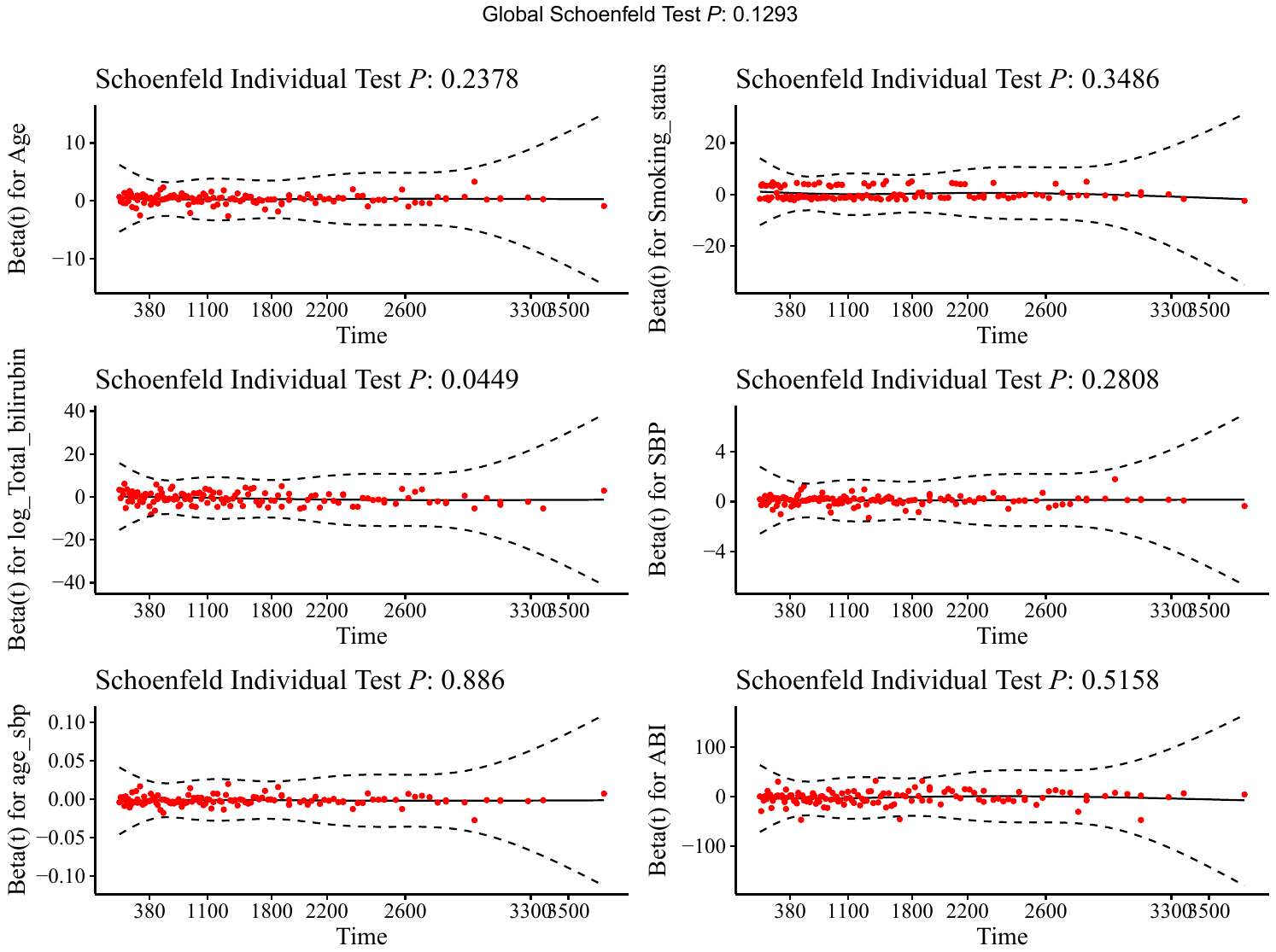

**Figure S1. The plots of the scaled Schoenfeld residuals against time in CVDMCM.**

**Online Methods**

**Measurement of clinical variables**

**Lifestyle variables**

Lifestyle variables included age, BMI, drinking, and smoking. BMI was computed as weight in kilograms divided by the square of height in meters. Drinking status was divided into two levels: 1) non-drinkers (people who never drink or who drink less than once a month, and alcohol content less than 10%); 2) drinkers (people who drink equal or more than once a month or alcohol content more than 10%). Smoking status was divided into three levels: 1) non-smokers (people who have never smoked before) or past smokers (people who used to smoke but have now quit); 2) current smokers (people who still smoke).

**Medical history**

Three medical history records, including hyperlipidemia, hypertension, and diabetes mellitus were obtained from in-person interviews. The diagnosis of hyperlipidemia was based on the history of lipid-lowering drug use or any one of the following: 1) LDL cholesterol (LDL-C) concentration of ≥4.14 mmol/L; 2) total cholesterol concentration of >6.45 mmol/L; 3) TG concentration of ≥2.26 mmol/L. The diagnosis of hypertension was based on resting blood pressure and a history of antihypertensive drug use. If the resting blood pressure was >140/90 mmHg, or with a history of antihypertensive drug use, the individual was diagnosed with hypertension. The diagnosis of diabetes mellitus was based on the history of antidiabetics drug use or any one of the following: 1) fasting plasma glucose concentration of ≥7.0 mmol/L; 2) plasma glucose concentration of ≥11.1 mmol/L 2 hours after a 75-g oral glucose load in a glucose tolerance test; 3) symptoms of high blood sugar and a casual plasma glucose concentration of ≥11.1 mmol/L; 4) HbA1c of ≥48 mmol/mol.

**Physical examination**

Blood pressure, baPWV, and ABI were measured using the Vascular Profiler BP-203RPEIII (Omron, Kyoto, Japan). The examination room was maintained at a standardized temperature of approximately 26℃. Trained technicians placed four pressure cuffs on the subjects (one on the upper part of each arm and one on each ankle). Then subjects were examined after 10 minutes of rest in the supine position. The device simultaneously recorded bilateral systolic and diastolic blood pressure, ABI, and baPWV, the latter of which was calculated as the ratio of travelled distance (which was automatically estimated from body height) divided by the transit time of the pulse wave between the brachial and posterior tibial arteries. The average of two-sided baPWV values and two-sided ABI values were recorded for analysis.

**Blood examination**

Blood examination, including routine blood tests and blood biochemical index tests, were measured using fasting venous blood samples. Routine blood tests were performed using the XN9000 (Sysmex, Kobe, Japan). Blood biochemical indices, including liver function, renal function, blood lipid profile, fasting blood glucose, HbA1c, and uric acid, were measured using the COBAS 8000 c701 (ROCHE, Basel, Switzerland).

**Urine examination**

Urinary elements were measured using the UF-1000i fully automatic urine analyzer (Sysmex, Kobe, Japan). Urinary chemistry elements were measured using the Siemens Atlas Urine Chemistry Analyzer (Siemens, Erlangen, Germany). The estimated glomerular filtration rate was calculated according to the Cockcroft–Gault formula.

**The online calculator**

We developed an online calculator for physicians to conveniently input patient data and generate CVD risk scores. The online calculator can be found on the website:

https://ctan2020.github.io/-calCVDrisk-/
